# Development of a bedside score to predict dengue severity

**DOI:** 10.1101/2020.11.25.20238972

**Authors:** Ingrid Marois, Carole Forfait, Catherine Inizan, Elise Klement-Frutos, Anabelle Valiame, Daina Aubert, Ann-Claire Gourinat, Sylvie Laumond, Emilie Barsac, Jean-Paul Grangeon, Cécile Cazorla, Audrey Merlet, Arnaud Tarantola, Myrielle Dupont-Rouzeyrol, Elodie Descloux

## Abstract

**Background:** In 2017, New Caledonia experienced an outbreak of severe dengue causing high hospital burden (4,379 cases, 416 hospital admissions, 15 deaths). We decided to build a local operational model predictive of dengue severity, which was needed to ease the healthcare circuit.

**Methods:** We retrospectively analyzed clinical and biological parameters associated with severe dengue in the cohort of patients hospitalized at the Territorial Hospital between January and July 2017 with confirmed dengue, in order to elaborate a comprehensive patient’s score. Patients were compared in univariate and multivariate analyses. Predictive models for severity were built using a descending step-wise method.

**Results:** Out of 383 included patients, 130 (34%) developed severe dengue and 13 (3.4%) died. Major risk factors identified in univariate analysis were: age, comorbidities, presence of at least one alert sign, platelets count <30×10^9^/L, prothrombin time <60%, AST and/or ALT >10N, and previous dengue infection. Severity was not influenced by the infecting dengue serotype nor by previous Zika infection. Two models to predict dengue severity were built according to sex. Best models for females and males had respectively a median Area Under the Curve = 0.80 and 0.88, a sensitivity = 84.5% and 84.5%, a specificity = 78.6% and 95.5%, a positive predictive value = 63.3% and 92.9%, a negative predictive value = 92.8% and 91.3%. Models were secondarily validated on 130 patients hospitalized for dengue in 2018.

**Conclusion:** We built robust and efficient models to calculate a bedside score able to predict dengue severity in our setting. We propose the spreadsheet for dengue severity score calculations to health practitioners facing dengue outbreaks of enhanced severity in order to improve patients’ medical management and hospitalization flow.

## Background

Dengue fever is the most prevalent human arbovirosis and a major public health issue in tropical and sub-tropical countries with epidemic outbreaks (1, 2). Dengue viruses are subdivided in 4 serotypes (DENV-1 to -4). There is a lack of specific treatments, vector control measures regularly fail to prevent epidemics and safe preventive dengue vaccines are not widely available (3-5). While new prevention methods are being developed, clinical management strategies are of prime importance.

Dengue has a wide spectrum of clinical presentations usually starting by an abrupt onset of fever, malaise, skin rash, headache, anorexia/vomiting, diarrhea, and abdominal pain, often with unpredictable clinical evolution. Clinical outcome can vary from a self-limiting non-severe condition to a potentially lethal disease subsequent to a vascular permeability resulting in leakage of fluids into serosal cavities and shock, hemorrhages, and/or organ failures (4). Warning signs of severe dengue include persisting vomiting, abdominal pain, lethargy/anxiety, mucosal bleeding, liquid accumulation, hepatomegaly, and rapid hematocrit increase concurrent with a platelet count drop (4).

In New Caledonia (NC), a French South Pacific Island Territory of 270,000 inhabitants, dengue is a closely monitored notifiable disease, enabling the collection of reliable documentation of dengue cases. Dengue fever outbreaks frequency is increasing in NC (**Fig 1**), and is associated to the emergent co-circulation of several DENV serotypes and other arboviruses, i.e. chikungunya and Zika viruses (6-8). An uninterrupted circulation of DENV-1 has been documented in NC between 2007 and 2018.

**Figure 1.**
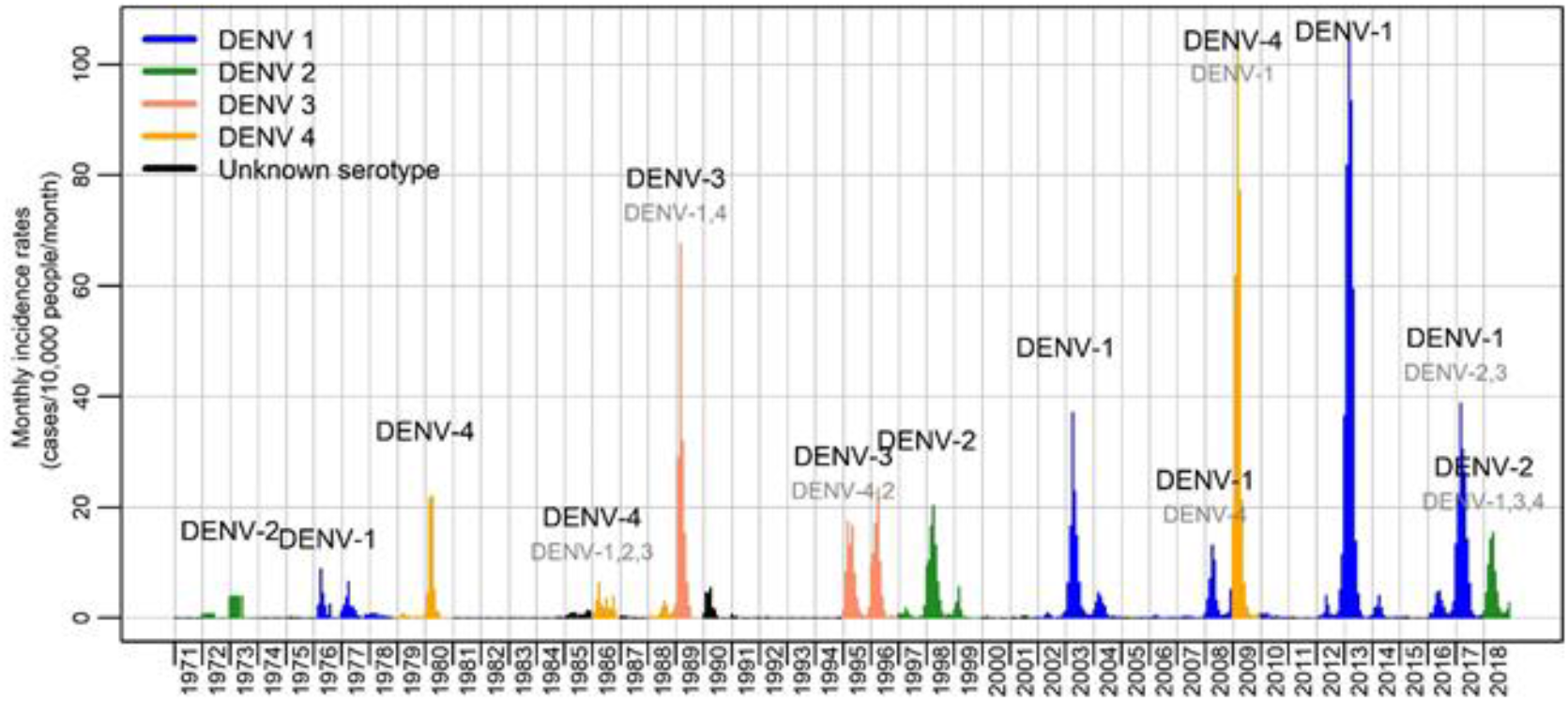
Monthly dengue incidence rates in New Caledonia from 1971 to 2018. Timeline of monthly dengue incidence rates per 10,000 inhabitants in New Caledonia from 1971 to 2018, showing an increasing frequency of outbreaks. For the first time in 2017, we observed a co-circulation of 3 serotypes (DENV-1, DENV-2, DENV-3).

During the 2017 dengue outbreak, three serotypes have co-circulated (DENV-1, DENV-2, DENV-3) for the first time. 4,379 dengue cases were declared among which 2,372 (54%) were biologically confirmed by RT-qPCR (6). Fifteen patients died (lethality rate= 0.3%). The hospitalization rate was exceptionally high (11.5% versus 3.5% during the 2012-2013 outbreak, 2.1% during the 2008-2009 outbreak, and 4.5% during the 2003 outbreak).

Identifying risk factors for severe dengue is of prime importance to improve patients’ medical care and better manage in-hospital patient flow. Such risk factors may differ depending on the region of the world considered, in link with populations’ genetics and way of living. To our knowledge, risk factors for severe dengue have been mostly explored in countries where dengue is endemic and have never been explored in the Pacific region, where dengue has an epidemic mode of circulation. Assessing the reliability of identified risk factor for severe dengue in epidemiological contexts where dengue has an epidemic mode of circulation is important to relieve the health care system upon outbreaks. Furthermore, the expansion of dengue in more temperate countries will certainly lead the hospitals to be overwhelmed, and operational tools developed in epidemic countries would help better manage in-hospital patient flow. In 2014-2015, the Pacific region and South America experienced a Zika pandemic. This pandemic occurred in countries where dengue circulates actively, it is therefore crucial to determine whether a previous Zika infection represents a risk factor for severe dengue.

The purposes of this study were to investigate clinical and biological parameters associated with severe dengue and elaborate an operational model to score patients’ risk to develop severe dengue in the NC medical facilities setting.

## Methods

### Study population

A total of 416 patients were admitted to the Territorial Hospital of New Caledonia between January 1^st^ 2017 and July 31^st^ 2017 with a diagnosis of dengue fever. Among them, 385 were biologically confirmed using RT-qPCR (9), of which 383 were enrolled in this study (**Supplementary Fig1**).

### Data collection

Patients’ clinical and biological characteristics were retrieved from hospital medical records (DxCare-Medasys), dengue notification sheets and completed by a telephone interview using a standardized questionnaire. Collected data included patients’ gender, age, ethnicity, medical history, treatments, substance abuse (tobacco, alcohol >3 units/day, cannabis, kava), clinical and biological parameters, presence of warning and severe signs, infecting dengue serotype and previous dengue and Zika infection. Dengue serotyping by RT-qPCR and IgG serology for dengue (PanBio®) and Zika (Euroimmun®) were performed.

### Patients Classification

Disease severity was assessed according to the WHO 2009 criteria (4). Patients were classified as severe when they displayed at least one of the following criteria: severe plasma leakage (shock, liquid accumulation visualized by sonogram or x-ray) with respiratory distress; severe hemorrhage; and/or severe organ failure (kidney, central nervous system, liver, heart). Thrombocytopenia with platelet count under 10×10^9^/L (reference range 150-400×10^9^/L) associated to minor bleeding was used as an additional severity criterion. Acute renal failure was defined as a Glomerular Filtration Rate by the MDRD Equation <60mL/min/1.73m^2^ or, for patients with previous chronic renal failure, a 2-fold increase from their baseline creatinine level. Severe hepatitis was defined by a transaminase (AST or ALT) level above 1000IU/L.

### Statistical Analysis and Predictive Models construction

For each quantitative variable, minimum, maximum, mean and median were calculated. Quantitative variables age and biological parameters were categorized into qualitative variables. Non-severe patients were compared to severe patients. In the univariate analysis, Odds Ratio (OR) and 95% confidence interval were calculated. Test of independence *p*-values were estimated using Fisher’s exact test. Differences were considered significant if *p*<0.05. Parameters for which Fisher test *p*≤0.2 were used to perform the multivariate analysis. Odds ratio were adjusted for each variable category with the value of the other variables being fixed. A predictive model for severe dengue was built using multiple logistic regression and a descending stepwise analysis. Statistics were performed using R software (version 3.5.1 (2018-07-02)).

### Predictive models validation

A k-fold cross-validation procedure (k=10) was used. The subsample k was retained as validation data and the remaining k-1 subsamples were used as training data. The cross-validation process was repeated k times, each of the k subsamples used exactly once as the validation data. Model performance was measured using the following indicators: sensitivity, specificity, positive predictive value, negative predictive value, Yule index (10), Youden index (sensitivity + specificity – 1) [(11) and the Area Under Curve (AUC) of the Receiving Operating Characteristic (ROC) curve. Using the variable coefficients determined in the logistic regression, patients’ score was expressed as p (probability to develop a severe dengue). The decision-making threshold was defined using the ROC curve as the best combination of sensitivity and specificity.

### Ethics Statement

Ethical approval was granted by the Consultative Ethics Committee of NC, and by the internal ethical review board of the Territorial Hospital. Dengue fever is a compulsory declarative disease in NC. Oral informed consent was obtained from all participating patients or their relatives retrospectively when consulted by telephone.

## Results

### Characteristics of the studied population

The characteristics of the 383 PCR-confirmed dengue patients included in this study are presented in **Table 1**. Patients were hospitalized on average on the 5^th^ day after symptom onset (median = 4, IQR = 3), for a median duration of 4 days (IQR = 3). They were 174 men and 209 women (Sex-ratio=0.83) with an age ranging from birth to 96 years old (IQR = 34, median 32 years). Symptoms and biological parameters available at hospital admission are summarized in **Table 2**. DENV-1 was the major serotype (80.6%), followed by DENV-2 (15.9%) and DENV-3 (3.6%). According to the WHO 2009 classification, 299/383 patients (78%) displayed at least one warning sign. Overall, 130 patients (34%) developed severe dengue.

**Table 1.** Characteristics of 383 hospitalized patients during the 2017 dengue outbreak in New Caledonia and results of the univariate analysis.

| Characteristics | Number (%) | Non severe cases (%)<br>n = 253 | Severe cases (%)<br>n = 130 | Odds ratio [CI 95%]<br>p value |
| --- | --- | --- | --- | --- |
| **Sex** |  |  |  |  |
| Men | 174 (45.4%) | 109 (43.1%) | 65 (50%) | 1.32 [0.86-2.02]<br>p=0.23 |
| Women | 209 (54.6%) | 144 (56.9%) | 65 (50%) | Reference |
| **Age class** |  |  |  |  |
| < 10 years old | 48 (12.4%) | 39 (15.4%) | 9 (6.9%) | 0.77 [0.28-2.06]<br>p=0.63 |
| ]10-20] | 69 (18.0%) | 50 (19.8%) | 19 (14.6%) | 1.26 [0.55-2.98]<br>p=0.68 |
| ]20-30] | 72 (18.8%) | 43 (17%) | 29 (22.3%) | 2.22 [1.01-5.11]<br>p=0.05 |
| ]30-40] | 52 (13.6%) | 40 (15.8%) | 12 (9.2%) | Reference |
| ]40-50] | 42 (11.0%) | 26 (10.3%) | 16 (12.3%) | 2.03 [0.83-5.11]<br>p=0.17 |
| ]50-60] | 39 (10.2%) | 24 (9.5%) | 15 (11.5%) | 2.06 [0.82-5.26]<br>p=0.16 |
| ]60-70] | 24 (6.3%) | 12 (4.7%) | 12 (9.2%) | 3.26 [1.16-9.44]<br>p=0.03 |
| > 70 | 37 (9.7%) | 19 (7.5%) | 18 (13.8%) | 3.10 [1.25-7.97]<br>p=0.02 |
| **Self-declared ethnicity** |  |  |  |  |
| Melanesian | 141 (36.7%) | 92(36.4%) | 49 (37.7%) | 1.30 [0.73-2.34]<br>p=0.37 |
| European | 86 (22.5%) | 61 (24.1%) | 25 (19.2%) | Reference |
| Polynesian | 68 (17.8%) | 43 (17%) | 25 (19.2%) | 1.40 [0.71-2.81]<br>p=0.39 |
| Métis /Other | 63 (16.4%) | 44(17.3%) | 19(14.6%) | 1.05 [0.51-2.15] p=1 |
| Not specified | 25 (5.1%) | 12 (9.2%) | 5 (3.8%) | 2.23[0.88-5.66] p=0.09 |
| **Risky behavior** |  |  |  |  |
| Tobacco | 105 (27.4%) | 62 (24.5%) | 43 (33.1%) | 1.52 [0.95-2.42]<br>*p*=0.09 |
| Cannabis | 19 (4.9%) | 12 (4.7%) | 7 (5.4%) | 1.15 [0.41- 2.97]<br>*p*=0.81 |
| Kava<sup>a</sup> | 15 (3.9%) | 10 (3.9%) | 5 (3.8%) | 0.98 [0.29-2.89] *p*=1 |
| Alcohol (>3 units/day) | 9 (2.3%) | 4 (1.6%) | 5 (3.8%) | 2.46 [0.62-10.56]<br>*p*=0.17 |
| ***Comorbidities*** |  |  |  |  |
| Obesity | 95 (24.8%) | 52 (20.6%) | 43 (33.1%) | 1.91 [1.18-3.07]<br>*p*=0.08 |
| Diabetes | 34 (8.8%) | 20 (7.9%) | 14 (10.8%) | 1.41 [0.67-2.89]<br>*p*=0.35 |
| Dyslipidemia | 27 (7.0%) | 13 (5.1%) | 14 (10.8%) | 2.22 [1.0-4.97] *p*=0.06 |
| Hypertension | 66 (17.2%) | 31 (12.3%) | 35 (26.9%) | 2.63 [1.53-4.54]<br>*p*<0.01 |
| Heart disease | 21 (5.5%) | 11 (4.3%) | 10 (7.7%) | 1.83 [0.74-4.51]<br>*p*=0.23 |
| Lung diseases | 31 (8.1%) | 24 (9.5%) | 7 (5.4%) | 0.55 [0.21-1.26]<br>*p*=0.23 |
| Renal failure | 9 (2.3%) | 4 (1.6%) | 5 (3.8%) | 2.46 [0.62-10.56]<br>*p*=0.17 |
| Immunodepression | 7 (1.8%) | 5 (2%) | 2 (1.5%) | 0.81 [0.10- 3.99] *p*=1 |
| Cancer | 14 (3.7%) | 8 (3.2%) | 6 (4.6%) | 1.49 [0.47-4.46]<br>*p*=0.57 |
| Risk of bleeding<sup>b</sup> | 9 (2.3%) | 6 (2.4%) | 3 (2.3%) | 1.0 [0.20-3.97] *p*=1 |
| ***Number of clinical problems*** |  |  |  |  |
| No medical history | 201 (52.5%) | 142 (56.1%) | 59 (45.4%) | Reference |
| 1 | 87 (22.7%) | 59 (23.3%) | 28 (21.5%) | 1.04 [0.58-1.80] *p*=1 |
| 2 or more | 95 (24.8%) | 52 (20.6%) | 43 (33.1%) | 2.12 [1.29-3.49]<br>*p*<0.01 |
| ***History of arbovirus infection*** |  |  |  |  |
| Presence of dengue IgG | 132 (34.5%) | 68 (26.9%) | 64 (49.2%) | 2.93 [1.83-4.75]<br>*p*<0.001 |
| Presence of Zika IgG | 42 (11.0%) | 25 (9.9%) | 17 (13.1%) | 1.30 [0.66-2.53]<br>*p*=0.49 |
| ***Treatment*** |  |  |  |  |
| Paracetamol | 300 (78.3%) | 203 (80.2%) | 97 (74.6%) | 0.72 [0.44-1.20]<br>*p*=0.24 |
| Corticosteroids | 2 (0.5%) | 0 | 2 (1.5%) | - |
| NSAI | 6 (1.6%) | 4 (1.6%) | 2 (1.5%) | 1.00 [0.12- 5.56] $p=1$ |
| Anticoagulants | 9 (2.3%) | 3 (1.2%) | 6 (4.62) | 3.92 [0.98-19.96]<br>$p=0.07$ |
| PAI | 38 (9.9%) | 19 (7.5%) | 19 (14.6%) | 2.10 [1.06- 4.17]<br>$p=0.03$ |
| Traditional medicine | 71 (18.5%) | 46 (18.2%) | 25 (19.2%) | 1.07 [0.62- 1.83]<br>$p=0.89$ |
| **Symptoms** |  |  |  |  |
| Fever | 347 (90.6%) | 231 (91.3%) | 116 (89.2%) | 0.79 [0.39-1.64]<br>$p=0.58$ |
| Muscle soreness/myalgia | 249 (65.0%) | 161 (63.6%) | 88 (67.7%) | 1.20 [0.77-1.88]<br>$p=0.50$ |
| Arthralgia | 174 (45.4%) | 110 (43.5%) | 64 (49.2%) | 1.26 [0.82-1.93]<br>$p=0.33$ |
| Headaches | 261 (68.1%) | 175 (69.2%) | 86 (66.2%) | 0.87 [0.56-1.37]<br>$p=0.56$ |
| Retro-orbital pain | 98 (25.6%) | 71 (28.1%) | 27 (20.8%) | 0.67 [0.40-1.11] |
| Diarrhea | 129 (33.7%) | 85 (33.6%) | 44 (33.8%) | 1.01 [0.64-1.58]<br>$p=1.0$ |
| Nausea/vomiting | 209 (54.6%) | 135 (53.4%) | 74 (56.9%) | 1.15 [0.75-1.77]<br>$p=0.52$ |
| Skin rash | 130 (33.9%) | 95 (37.5%) | 35 (26.9%) | 0.61 [0.38-0.97]<br>$p=0.041$ |
| Conjunctival hyperemia | 41 (10.7%) | 33 (13.0%) | 8 (6.2%) | 0.44 [0.18-0.95]<br>$p=0.053$ |
| Edema | 18 (4.7%) | 8 (3.2%) | 10 (7.7%) | 2.54 [0.96-6.90]<br>$p=0.071$ |
| Gingivorrhagia | 58 (15.1%) | 33 (13.0%) | 25 (19.2%) | 1.59 [0.89-2.80]<br>$p=0.13$ |
| Purpura | 78 (20.4%) | 47 (18.6%) | 31 (23.8%) | 1.37 [0.81-2.29]<br>$p=0.23$ |
| Epistaxis | 58 (15.1%) | 34 (13.4%) | 24 (18.5%) | 1.46 [0.81-2.58]<br>$p=0.23$ |
| Hematuria/blood in stools | 19 (5.0%) | 6 (2.4%) | 13 (10%) | 4.49 [1.71-13.30]<br>$p<0.01$ |
| **Alert signs** |  |  |  |  |
| Abdominal pain | 147 (38.4%) | 88 (34.8%) | 59 (45.4%) | 1.56 [1.01-2.40]<br>$p=0.046$ |
| Persistent vomiting | 42 (11.0%) | 28 (11.1%) | 14 (10.7%) | 0.97 [0.48-1.90] $p=1.0$ |
| Clinical liquid accumulation | 28 (7.3%) | 11 (4.3%) | 17 (13.1%) | 3.28 [1.50-7.50]<br>$p<0.01$ |
| Mucosal bleeding | 170 (44.4%) | 83 (32.8%) | 87 (66.9%) | 4.12 [2.64-6.51]<br>*p*<0.01 |
| Lethargy/anxiety | 72 (18.8%) | 46 (18.2%) | 26 (20.0%) | 1.13 [0.65-1.92]<br>*p*=0.68 |
| Hepatomegaly | 17 (4.4%) | 9 (3.6%) | 8 (6.2%) | 1.78 [0.64-4.83]<br>*p*=0.30 |
| Increase in Ht and platelet count drop | 68 (17.8%) | 37 (14.6%) | 31 (23.8%) | 1.83 [1.07-3.12]<br>*p*=0.034 |
| **Biological parameters** |  |  |  |  |
| Normal platelet count | 225 (58.7%) | 175 (69.1%) | 50 (38.4%) | Reference |
| Platelets < 30x10<sup>9</sup>/L | 134 (35.0%) | 59 (23.3%) | 75 (57.7%) | 4.42 [2.79–7.08]<br>*p*<0.01 |
| GFR < 60 ml/min | 42 (11.0%) | 8 (3.1%) | 34 (26.1%) | 9.28 [4.22–22.90]<br>*p*<0.01 |
| Normal AST | 217 (56.7%) | 157 (62%) | 60 (46.1%) | Reference |
| AST > 10N | 110 (28.7%) | 51 (20%) | 59 (45.4%) | 3.01 [1.87–4.89]<br>*p*<0.01 |
| Normal ALT | 275 (71.8%) | 198 (78.3%) | 77 (59.2%) | Reference |
| ALT > 10N | 51 (13.3%) | 10 (3.9%) | 41 (31.5%) | 10.34 [5.10–22.94]<br>*p*<0.01 |

<sup>a</sup>Kava: traditional beverage produced from *poivrier* roots, consumed throughout the cultures of Polynesia, Melanesia, and parts of Micronesia for its sedating and euphoriant effect.
<sup>b</sup>Risk of bleeding refers to comorbidities with previous risk of hemorrhage (menorrhagia, endometriosis, adenomyosis, gastric ulcer, immunological thrombopenic purpura).
PAI = Platelet Aggregation Inhibitor, NSAID = Non-Steroidal Anti-Inflammatory, GFR = Glomerular Filtration Rate, AST: Aspartate AminoTransferase, ALT: Alanine AminoTransferase.

**Table 2.** Clinical and biological parameters at hospital admission in the 383 hospitalized patients.

| Clinical parameters | Number (%) |
| --- | --- |
| **Symptom** |  |
| Fever | 347 (90.6%) |
| Muscle soreness/myalgia | 249 (65.0%) |
| Arthralgia | 174 (45.4%) |
| Headache | 261 (68.1%) |
| Retro-orbital pain | 98 (25.6%) |
| Diarrhea | 129 (33.7%) |
| Nausea/vomiting | 209 (54.6%) |
| Skin rash | 130 (33.9%) |
| Conjunctival hyperemia | 41 (10.7%) |
| Edema | 18 (4.7%) |
| Gingivorrhagia | 58 (15.1%) |
| Purpura | 78 (20.4%) |
| Epistaxis | 58 (15.1%) |
| Hematuria/blood in stools | 19 (5.0%) |
| Shock syndrome | 26 (6.8 %) |
| Major bleeding | 47 (12.3%) |
| **Alert signs** |  |
| Abdominal pain | 147 (38.4%) |
| Persistent vomiting | 42 (11.0%) |
| Clinical liquid accumulation | 28 (7.3%) |
| Mucosal bleeding | 170 (44.4%) |
| Lethargy/anxiety | 72 (18.8%) |
| Hepatomegaly | 17 (4.4%) |
| Increase in hematocrit + drop in platelets count | 68 (17.8%) |
| **Biological parameters** | **Median [min; max]**<br>**(% of available results)** |
| Platelets (10<sup>9</sup>/L) | 48 [3; 360](93.7) |
| Hemoglobin (g/dL) | 14 [6; 22] (94.5) |
| Hematocrit (%) | 41 [16; 61] (94) |
| Neutrophils (/mm<sup>3</sup>) | 1,895 [320; 19,020] (92) |
| Lymphocytes (/mm<sup>3</sup>) | 1,535 [140; 8,940] (90.8) |
| Albuminaemia (g/L) | 36 [19; 46] (13.8) |
| Protidaemia (g/L) | 58 [24; 91] (10.4) |
| Urea (mmol/L) | 4 [0; 41](76) |
| Creatinin (μmol/L) | 71 [18; 927] (78.3) |
| AST (IU/L) | 184 [17; 10,336](85) |
| ALT (IU/L) | 116 [9; 8,040] (85) |
| CPK | 305 [3- 74,063] (28) |
| Lipase (IU/L) | 53 [9; 2,707](33.4) |
| CRP (mg/L) | 14 [0; 327] (56.6) |

While 121 patients (40%) with at least one warning sign developed severe dengue, only nine patients (10.7%) without warning signs did so (**Fig 2**). The Odds ratio (OR) for patients presenting at least one warning sign is 5.6 [2.17; 13.3], with a corresponding Relative Risk of 3.8 [2.0; 7.1]. Hepatitis was the most frequent severity criterion accounting for 41.5% of severe cases. Twenty-two patients (5.7%) developed major hemorrhage, 47 (12.3%) had deep thrombocytopenia <10×10^9^/L with minor bleeding, and 26 (6.8%) developed a shock, corresponding to 16.9%, 36.2% and 20% of severe cases, respectively (**Fig 3**). A total of 182 (47.5%) patients presented comorbidities. The most frequent comorbidity was obesity (24.8%), accounting for 33.1% of severe cases (**Table 1**). Thirteen patients died (lethality rate = 3.4%): ten during hospitalization and three after discharge. In-hospital deaths involved 7/10 patients younger than 55 years old without notable medical history. Previous dengue infection was recorded in 7/8 patients who died within a week; all three circulating serotypes were involved, including one case of coinfection.

**Figure 2.**
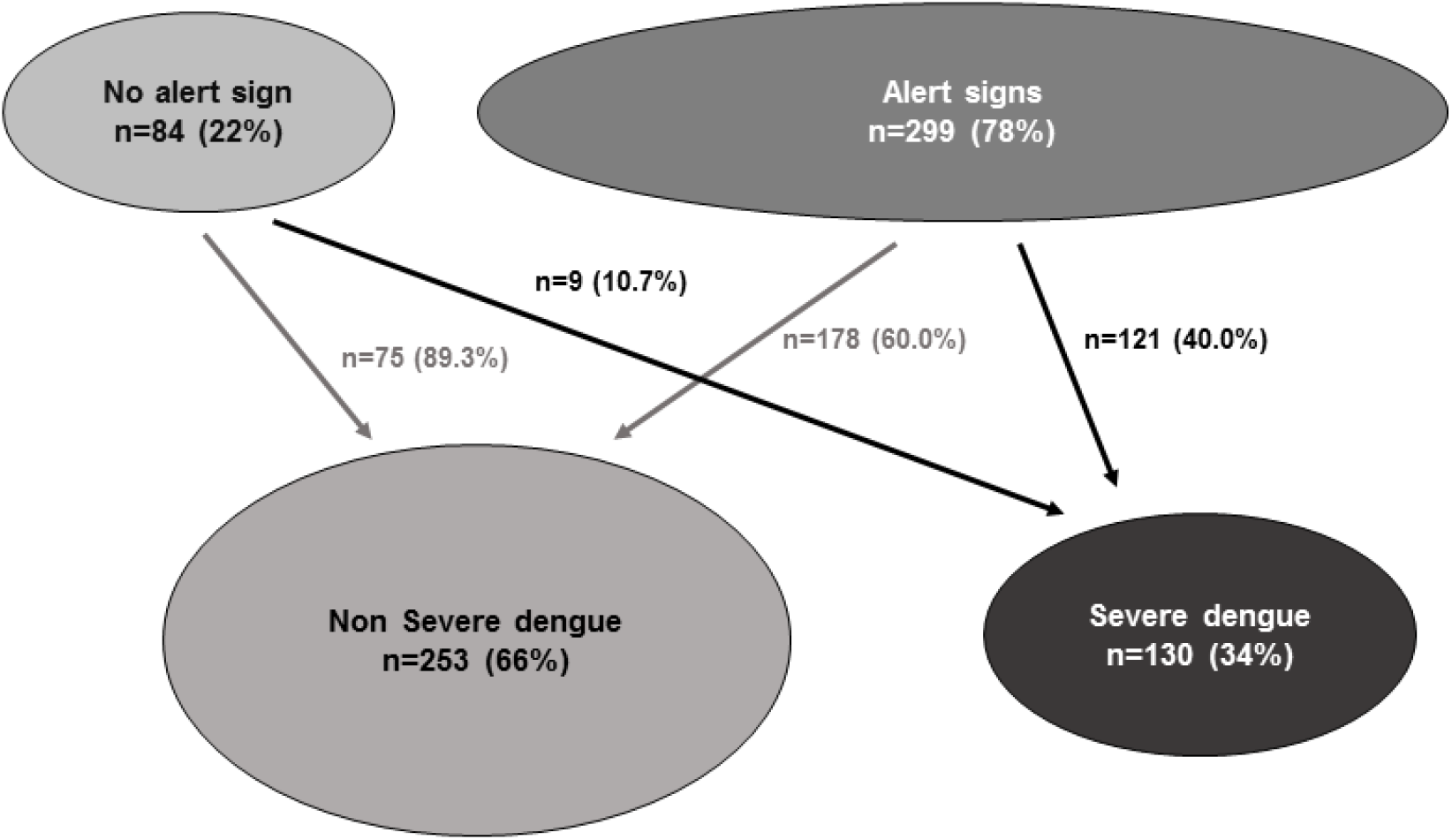
Classification of the 383 hospitalized patients according to the presence of alert and severity signs (2017 dengue outbreak, New Caledonia). Scheme of dengue cases distribution, showing the percentage of cases with and without alert signs and their evolution to non-severe and severe dengue, according to the WHO 2009 classification adapted for our study with minor modifications (thrombocytopenia <10×109/L associated to minor bleeding was used as an additional severity criterion).

**Figure 3.**
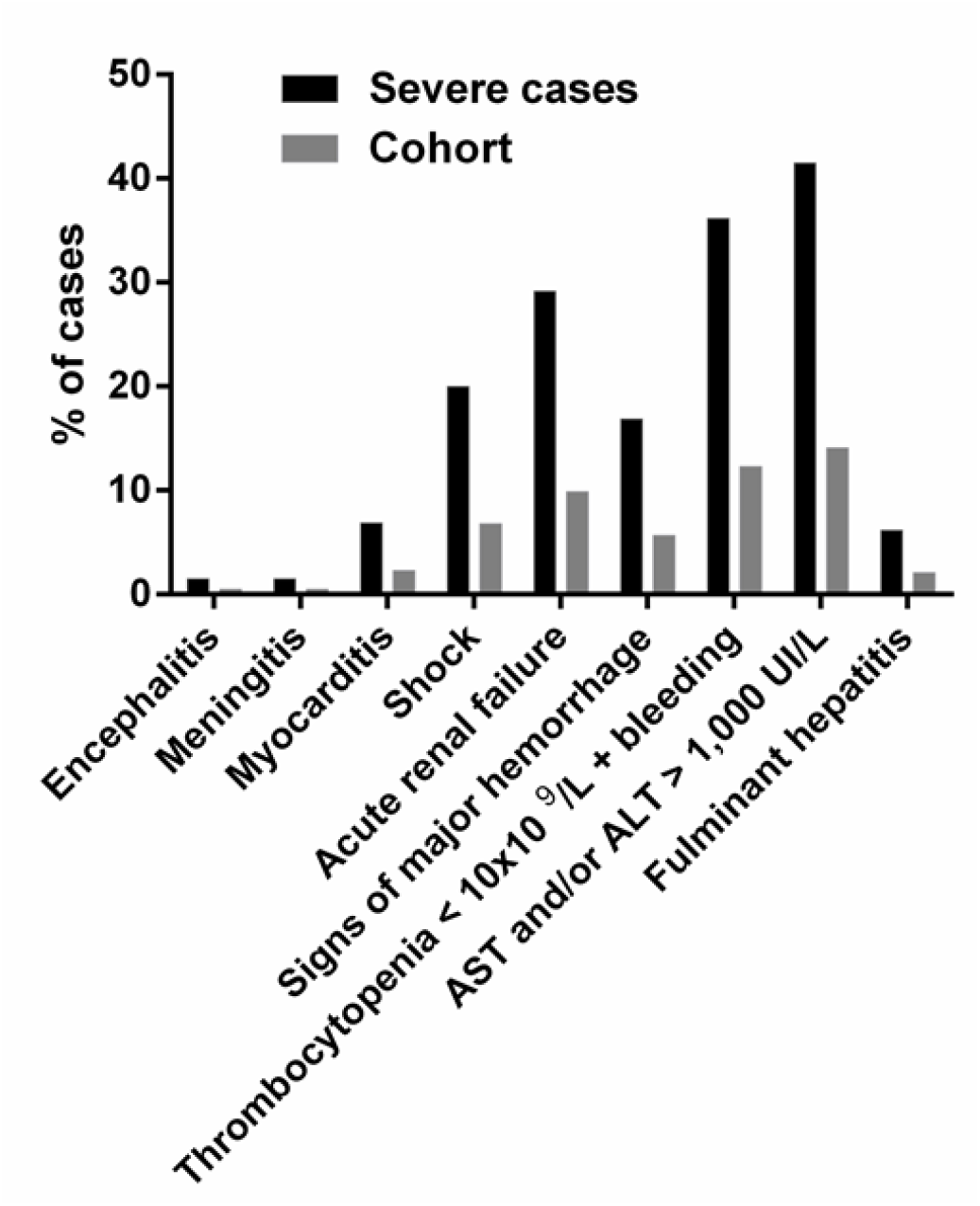
Clinical signs of severity and comorbidities. Percentage of cases exhibiting the indicated clinical signs of severity within the cohort (gray) and among severe cases (black).

### Factors associated with severe dengue in univariate analysis

Factors highly associated with severe dengue in univariate analysis included age [20-30] years and > 60 years (OR 2.22 and 3.26), comorbidities (hypertension OR 2.63, obesity OR 1.91, dyslipidaemia OR 2.22 and more than two comorbidities OR 1.98), previous dengue infection (OR 4.7), use of platelet aggregation inhibitors (OR 2.1), presence of at least one alert sign (OR 5.6) and prothrombin time < 60% (OR 7.2) (**Table 1**).

### Multivariate analysis and construction of the predictive models

Variables significantly associated with dengue severity in univariate analysis with *p*≤0.2 and available at hospital admission were taken into account in the multivariate analysis. These variables were: comorbidities (hypertension, myocardiopathy, dyslipidemia, obesity), excessive tobacco and alcohol consumption, anticoagulant or use of platelet aggregation inhibitors, mucosal bleeding, hematuria and/or presence of blood in stools, skin rash, clinical liquid accumulation, abdominal pain, simultaneous hematocrit increase and platelet count drop, platelets <30×10^9^/L, ALT and/or AST >10N. Raw analysis of the dataset showed that certain age groups in females were more at risk to develop severe dengue, while no difference was observed between age groups in men. This suggested the existence of an interaction between age and sex. This interaction was confirmed using a bivariate analysis taking age and sex as variables. Multivariate analysis further confirmed this interaction between age and sex: women between 20 and 30 years and men over 60 years were more at risk for severe dengue (OR 4.02 and 3.17, respectively). An analysis for each modality of the variable sex was thus performed and two different predictive models were built according to sex.

Descending stepwise analysis identified the best explanatory variables for progression towards severe dengue (**Table 3**). In the model for females, these variables were: age class, hypertension, skin rash, mucosal bleeding, platelets count <30×10^9^/L and ALT >10N. In the model for males, these variables were: age class, excessive alcohol consumption, mucosal bleeding, platelets count <30×10^9^/L and ALT >10N. The predictive models provide a score. Score threshold is derived from the ROC curve as the threshold optimal value for which sensitivity and specificity are the highest for the whole dataset. A score ≥ 0.36 for females and ≥ 0.34 for males indicates a high probability to develop severe dengue. The Excel spreadsheet enabling the calculation of this risk score is presented in **Supplementary Fig2**. Examples of models’ usage are presented in **Supplementary Fig3**.

**Table 3.** Results of multivariate analysis concerning determinant factors of dengue severity used to build the predictive models for females and males.

|  | Crude odds ratio | Adjusted odds ratio (females) | Adjusted odds ratio (males) |
| --- | --- | --- | --- |
| **Age class (years)** |  |  |  |
| ≤10 | 0.77 [0.28 – 2.05] | 2.52 [0.39 – 16.94] | 0.77 [0.12 – 4.72] |
| ]10-20] | 1.26 [0.55 – 2.98] | 3.22 [0.62 – 19.57] | 0.88 [0.18 – 4.52] |
| ]20-30] | 2.22 [1.01 – 5.10] | 7.79 [1.87 – 41.86] | 0.26 [0.04 – 1.57] |
| ]40-60] | 2.04 [0.94 – 4.64] | 5.74 [1.33 – 31.35] | 1.21 [0.24 – 6.37] |
| >60 | 3.17 [1.42 – 7.44] | 3.54 [0.58 – 24.36] | 8.45 [1.59 – 53.3] |
| Hypertension | 2.7 [1.6 – 4.7] | 4.68 [1.24 – 19.75] |  |
| Alcohol consumption | 2.46 [0.62 – 10.56] |  | 20.83 [1.93 – 807.49] |
| Mucosal bleeding | 4.12 [2.64 – 6.51] | 4.66 [2.08 – 11.14] | 9.79 [3.75 – 28.72] |
| Clinical liquid accumulation | 3.28 [1.50 – 7.50] | 3.88 [0.87 – 18.19] |  |
| Skin rash | 0.61 [0.38 – 0.97] | 0.41 [0.16 – 0.97] |  |
| **Platelets** |  |  |  |
| <30.10<sup>9</sup>/L | 4.42 [2.79 – 7.08] | 2.83 [1.26 – 6.45] | 5.84 [2.21 – 17.04] |
| **ALT (IU/L)** |  |  |  |
| ≥10N | 10.34 [5.10 – 22.94] | 14.31 [4.93 – 47.67] | 243.09 [28.75 – 6130.86] |

*All parameters are risk factors for dengue severity albeit skin rash that appears as a protective factor to develop severe dengue in females.*

### Performance of the models

A k-fold cross-validation procedure (k=10) showed that both models for females and males were robust and efficient (**Fig 4**), yielding a median AUC of 0.80 (Interquartile Range IQR = 0.08, range [0.638; 0.952]) and 0.88 (IQR=0.13, range [0.701; 1.00]), respectively and a high median Negative Predictive Value (92.8 and 91.3%, respectively). Models were challenged on a cohort of 130 patients (66 females, 64 males) hospitalized for dengue in NC in 2018. Contingency tables depicting the number of severe dengue cases among the 2018 dataset and the number of severe dengue retrieved from models predictions are shown in **Supplementary Fig4**. Models for females and males yielded a sensitivity of 73 and 84%, a specificity of 88 and 71%, a Positive Predictive Value of 65 and 55%, a Negative Predictive Value of 92 and 91%, a Youden index of 0.62 and 0.55 and a Yule Q index of 0.91 and 0.62, respectively.

**Figure 4.**
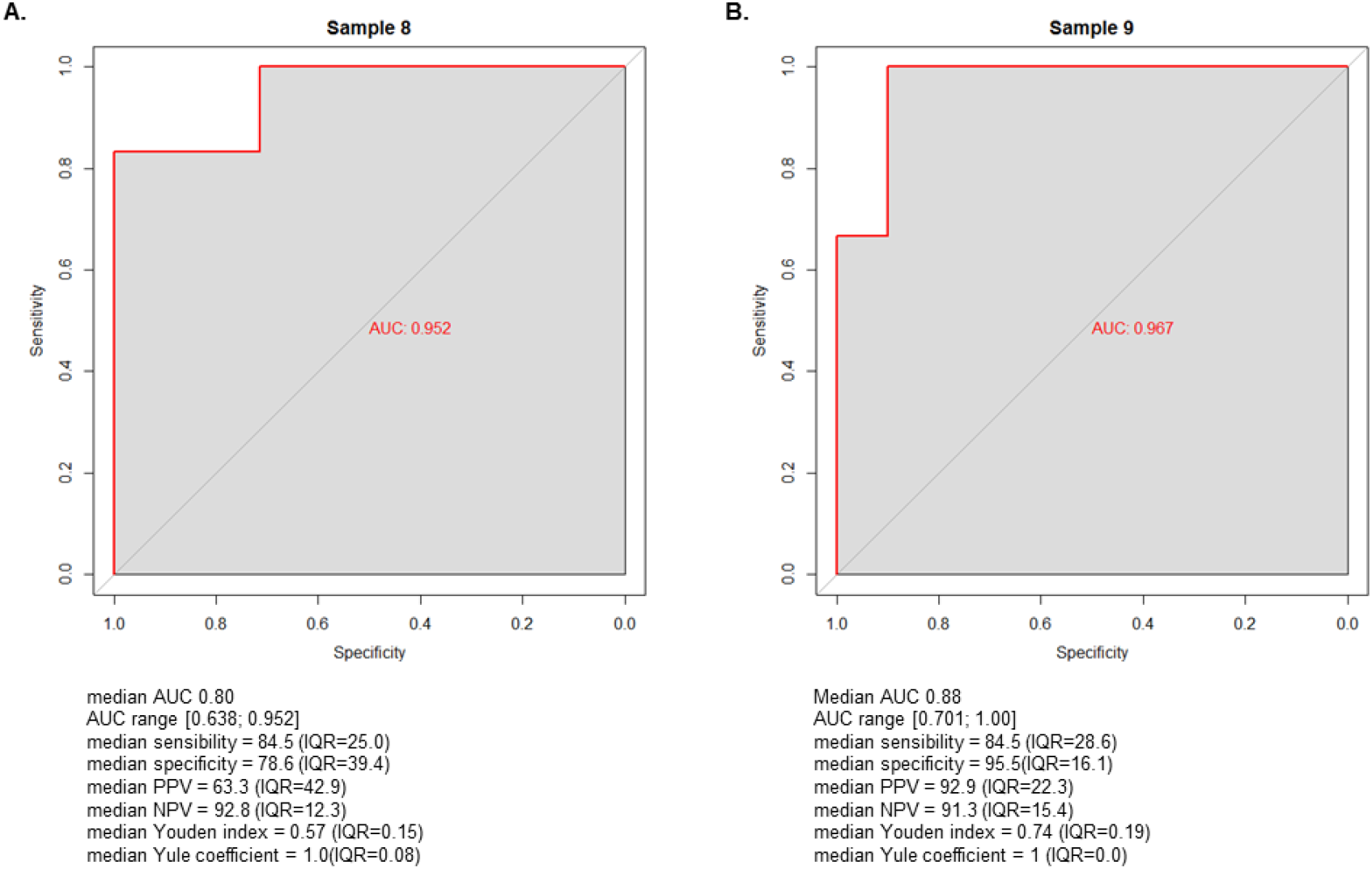
Performance of predictive models for severe dengue according to the sex, New Caledonia 2017. Receiving Operating Characteristic (ROC) curve for the best model for females (A) and the best model for males (B). Median AUC, Sensitivity, Specificity, Positive Predictive Value (PPV), Negative Predictive Value (NPV), Youden index and Yule Q coefficient are indicated for each model.

## Discussion

The 2017 dengue outbreak in New Caledonia was characterized by a high hospitalization rate leading to an overwhelming of emergency rooms and hospitalization units. Clinicians and epidemiologists expressed the necessity to develop a comprehensive operational tool in order to improve medical care and in-patients flow in local hospitals. Based on a detailed analysis of 383 hospitalized patients, we identified important demographical, clinical and biological parameters associated with severe dengue. Parameters easily available in routine practice were used to score patients’ risk of developing severe dengue.

Most of the risk factors we identified in univariate analysis have been described in other studies, i.e. age (12, 13), comorbidities such as hypertension (14) or diabetes (12, 14, 15), persistent vomiting (13, 16), increase in hematocrit (12, 16). Therefore, we confirm that the WHO 2009 criteria to evaluate the severity of dengue infection are applicable in NC. A platelet count <30×10^9^/L was also strongly associated with severe dengue in our study (OR 4.5) like in other studies (12, 16, 17). However, no consensus on severe thrombocytopenia definition was obtained in a recent working group of dengue researchers and public health specialists to develop standardized endpoints, and they remained divided on whether a rapid decreasing trend or a specific platelet count should be case-defining (18). Factors not associated with severity in NC are sex, ethnicity and substance abuse. In our study, the detected serotype responsible for the acute infection is not linked to the severity of the disease, unlike previous findings (19).

We report a 4.7 fold increased risk of severe dengue in patients who had a serologically documented history of previous dengue infection, in accordance with the Antibody-Dependent Enhancement (ADE) theory stating that secondary dengue infections are usually more severe (20). However, as dengue serology is usually not available at hospital admission, the presence of dengue IgG antibodies was not included in our multivariate analysis. For the first time to our knowledge, we have investigated the influence of a previous Zika infection on dengue outcome, and we found no association with dengue severity. As DENV and ZIKV serological tests display partial cross-reactivity (21), the impact of previous DENV and ZIKV infection on dengue severity should be confirmed by seroneutralization tests. We were able to build two robust and efficient logistic regression models to evaluate patients’ risk of developing severe dengue in men and women. These models enable the calculation of a risk score based on simple parameters and may represent easy-to-use operational tools to help clinicians in hospitalization decision and improve in-hospital patient flux (**Supplementary Fig 2&3**). Different predictive models have been proposed in previous studies, based on multivariate logistic regression (22-24) or on classification and regression trees (25, 26). Parameters used in these models were mainly age, leukocytosis and platelet count. Interestingly, Nguyen et al. developed a prognostic model taking into account vomiting, platelet count (< 10×10^9^/L), AST level (2-fold increase) and NS1 rapid test status (24). The model they propose yielded a very good discriminative ability (AUC 0.95), which is close to the AUC of 0.80 and 0.88 yielded by our models for females and males, respectively. However, a major caveat to their study is the absence of model cross-validation.

The bedside scoring tool we propose is very simple and easy to use, results ranging from 0 (minimal risk of severe dengue) to 1 (maximal risk). When a patient refers to the hospital with probable or confirmed dengue fever and has no other classical criteria of hospitalization, we calculate his severity score. If it is ≤0.34 for a man and ≤0.36 for a woman, we invite the patient to go back home and to be reassessed by a physician at day 4-6 post-symptoms’ onset. If the score is greater, we propose an admission for IV fluid treatment and bio-clinical monitoring. If the score is >0.6 the patient should be closely monitored and if it is >0.8 he might require intensive care. We also systematically recommend ICU admission when platelet count is <10×10^9^/L or transaminases >2000 IU/L.

Although based on a well-documented and validated database, biological data were heterogeneous as collected at different time points after symptoms’ onset, but this limit is inherent to retrospective studies. As our studied population was composed of relatively old patients (which is a characteristic of dengue epidemiology in NC), hospitalized in a single hospital (albeit the largest in NC), our models may not be valid to predict patients’ risk to develop severe dengue in other populations. However, our models were prospectively validated on 130 patients during 2018 DENV-2 outbreak in NC, yielding a high Negative Predictive Value of 92 and 91% for females and males respectively. Our models could thus be relevant for dengue severity prediction regardless of the incriminated serotype.

## Conclusions

We developed a bedside score to predict dengue severity. We propose this bedside score to be deployed and tested in other countries with similar dengue epidemiology, in order to optimize patients’ triage, in-hospital patients flux, and improve personalized medical care, thus benefiting both health practitioners and populations facing dengue outbreaks of enhanced severity.

## Data Availability

All data generated or analysed during this study are included in this published article.

## Abbreviations

ADE: Antibody-Dependent Enhancement
ALT: Alanine Aminotransferase
AST: Aspartate Aminotransferase
AUC: Area Under the Curve
DENV: dengue virus
ICU: Intensive Care Unit
IgG: type G Immunoglobulin
IQR: Interquartile Range
IU/L: International Units/L
NC: New Caledonia
NS1: Non-Structural protein 1
OR: Odds Ratio
ROC curve: Receiving Operating Characteristic curve
RT-qPCR: Reverse Transcription-quantitative Polymerase Chain Reaction
WHO: World Health Organization
ZIKV: Zika virus

## Declarations

### Ethics approval and consent to participate

Ethical approval was granted by the Consultative Ethics Committee of NC, and by the internal ethical review board of the Territorial Hospital. Dengue fever is a compulsory declarative disease in NC. Oral informed consent to participate was obtained from all participating patients or their relatives retrospectively when consulted by telephone.

### Consent to publish

Not applicable.

### Availability of data and materials

All data generated or analysed during this study are included in this published article

## Authors’ Contributions

ED, AV and CF conceived the study, IM, EK-F, CC, AM, ED recruited the patients. A-CG, EB performed the biological analyses. IM, CI, CF, AT, ED analyzed the data, CF performed the statistical analyses, AV, DA, SL, J-PG provided access to patients’ data. CI, AT, MD-R, IM, ED and EK-F wrote the manuscript, IM, CF, CI, EK-F and ED contributed equally to this work, all authors carefully revised the manuscript.

## Acknowledgements

We thank Sébastien Mabon, Sophie Lafleur, Pascal André, and Mathieu Serié from CHT Noumea for their help during this study, Antoine Biron, Marie-Amélie Goujart, Nathalie Amedeo, Erwan Choblet, Gauthier Delvallez from CHT Laboratory. We thank Ludovic Floury, Viktoria Taofifenua, Anne Pfannstiel from DASS-NC, Morgan Mangeas and Magali Teurali (IRD) for their help in statistics.

## Supplementary Material

**S1 Fig. STROBE flowchart describing patients enrolment in the study**.

**S2 Fig. Excel spreadsheet enabling the calculation of a bedside score predictive of severe dengue**. In the operating tool, scores derived from the logistic regression models can be calculated using an Excel spreadsheet by inserting 1 if the characteristic is present. A score ≥0.36 for females and 0.34 for males indicates a high probability to develop severe dengue. The hospitalization decision is made according to the medical opinion. During hospitalization, patients are submitted to close surveillance, hyperhydration, symptomatic treatments and sometimes blood support and resuscitation measures.

**S3 Fig. Examples of scoring to estimate the risk to develop severe dengue using data available at the moment of hospital admission decision**. In the upper example, the score of the female patient is above 0.36, indicating a high risk to develop severe dengue. In the lower example, the score of the male patient is below 0.34, indicating a low probability to develop severe dengue.

**S4 Fig. Contingency tables showing the performance of the models for the prediction of dengue severity on dengue 2018 outbreak in New Caledonia**. Absolute numbers of severe dengue observed in the dataset and predicted by the models are shown for the model for females (upper table) and the model for males (lower table).

