## Supplementary Material for "Development of a bedside score to predict dengue severity"

### Supporting information

**S1 Fig.**

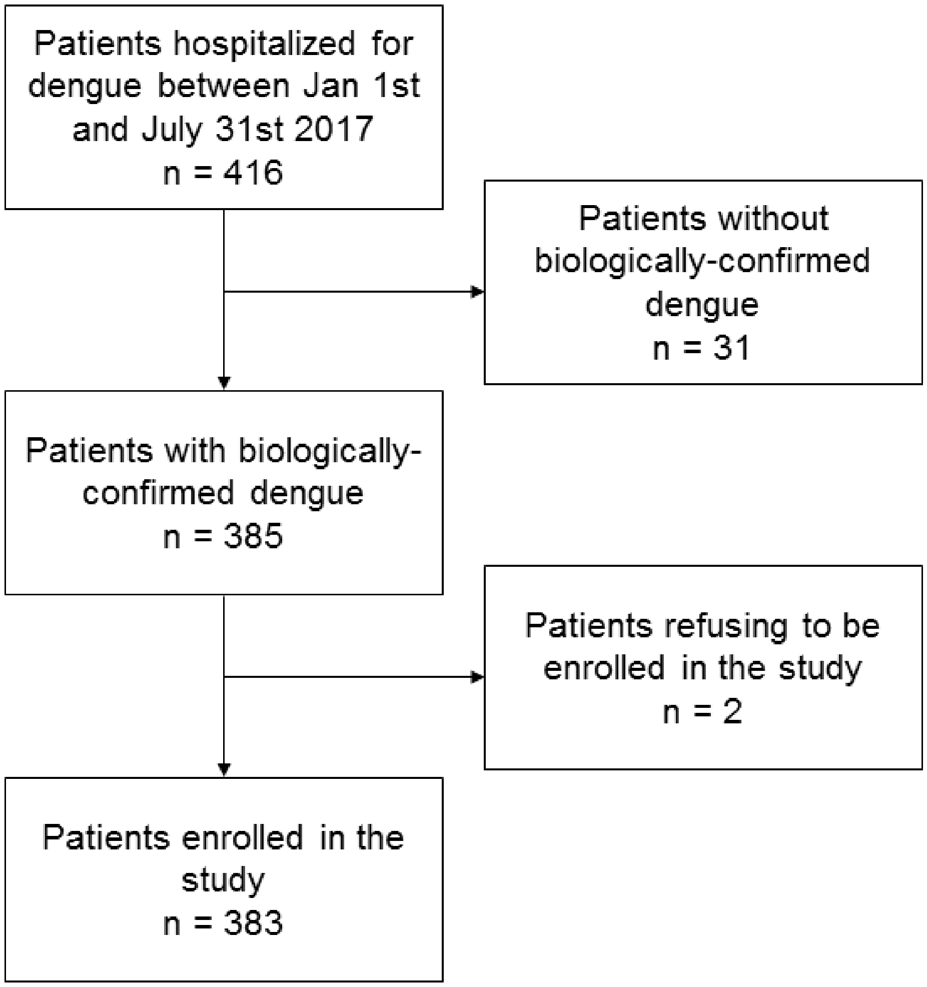

**S2 Fig.**

| Model for females | | If characteristic present, write 1 in the corresponding cell | Score |
|---|---|---|---|
| Age class (years) | >60 | | 0,02 |
| | ]40-60] | | |
| | ]30-40] | | |
| | ]20-30] | | |
| | ]10-20] | | |
| | <=10 | | |
| Medical history | Hypertension (treated or not) | | |
| Symptoms | Mucosal bleeding | | |
| | Clinical liquid accumulation | | |
| | Skin rash (except purpura) | | |
| Last biological results | Platelets <30 G/L | | |
| | ALT>10N | | |

| Model for males | | | |
|---|---|---|---|
| Age class (years) | >60 | | 0,04 |
| | ]40-60] | | |
| | ]30-40] | | |
| | ]20-30] | | |
| | ]10-20] | | |
| | <=10 | | |
| Risky behavior | Alcohol abuse >3 units/day | | |
| Symptoms | Mucosal bleeding | | |
| Last biological results | Platelets <30 G/L | | |
| | ALT>10N | | |

**S3 Fig.**

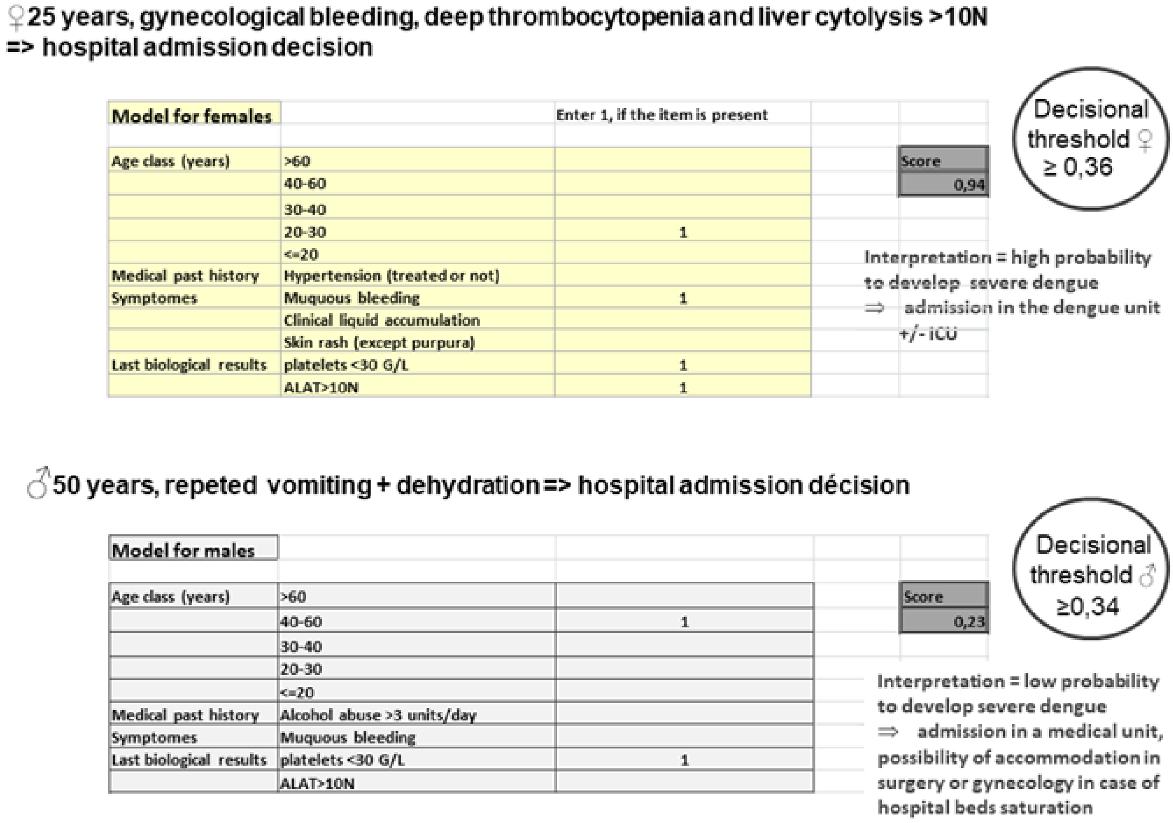

♀25 years, gynecological bleeding, deep thrombocytopenia and liver cytolysis >10N => hospital admission decision

| Model for females | | Enter 1, if the item is present |
|---|---|---|
| Age class (years) | >60 | |
| | 40-60 | |
| | 30-40 | |
| | 20-30 | 1 |
| | <=20 | |
| Medical past history | Hypertension (treated or not) | |
| Symptomes | Muquous bleeding | 1 |
| | Clinical liquid accumulation | |
| | Skin rash (except purpura) | |
| Last biological results | platelets <30 G/L | 1 |
| | ALAT>10N | 1 |

Score: 0,94

Decisional threshold ♀ ≥ 0,36

Interpretation = high probability to develop severe dengue
⇒ admission in the dengue unit +/- ICU

♂50 years, repeted vomiting + dehydration => hospital admission décision

| Model for males | | |
|---|---|---|
| Age class (years) | >60 | |
| | 40-60 | 1 |
| | 30-40 | |
| | 20-30 | |
| | <=20 | |
| Medical past history | Alcohol abuse >3 units/day | |
| Symptomes | Muquous bleeding | |
| Last biological results | platelets <30 G/L | 1 |
| | ALAT>10N | |

Score: 0,23

Decisional threshold ♂ ≥0,34

Interpretation = low probability to develop severe dengue
⇒ admission in a medical unit, possibility of accommodation in surgery or gynecology in case of hospital beds saturation

**S4 Fig.**

Females (n=66)

| Classification of the patients / Result of the model | Severe dengue | Non-severe dengue |
| --- | --- | --- |
| Severe dengue | 11 | 6 |
| Non-severe dengue | 4 | 45 |

Males (n=64)

| Classification of the patients / Result of the model | Severe dengue | Non-severe dengue |
| --- | --- | --- |
| Severe dengue | 16 | 13 |
| Non-severe dengue | 3 | 32 |
